# A National Study of Expedited Partner Therapy Use in Emergency Departments: A Survey of Medical Director Knowledge, Attitudes and Practices

**DOI:** 10.1101/2023.04.01.23287999

**Authors:** Rachel E Solnick, Rafael Cortes, Ethan Chang, Paul Dudas, Daxuan Deng, Cornelius Jamison, Okeoma Mmeje, Keith E. Kocher

**Author notes:** **Prior Presentations:** SAEM 2021 Virtual Conference. Author contributions: *Concept and design: Solnick, Kocher, Jamison, Mmeje, Acquisition, analysis, or interpretation of data: Solnick, Cortes, Dudas Drafting of the manuscript: Solnick, Chang, Cortes Critical revision of the manuscript: Solnick, Kocher, Jamison, Mmeje, Statistical analysis: Solnick, Deng Supervision: Solnick, Kocher. **Conflict of Interest Disclosure:** Authors report no conflict of interest.

## Abstract

**Background:** Emergency departments (EDs) are the primary source of healthcare for many patients diagnosed with sexually transmitted infections (STIs). Expedited partner therapy (EPT), treating the partner of patients with STIs without an exam of the partner, is an evidence-based practice for patients who might not otherwise seek care. Little is known about EPT use in the ED. In a national survey, we describe ED medical directors’ knowledge, attitudes, and practices of EPT.

**Methods:** A cross-sectional internet survey of medical directors from academic EDs using the Academy of Academic Administrators of Emergency Medicine (AAAEM) Benchmarking Group from July through September 2020. Primary outcomes were EPT awareness, support, and use. The survey also examined barriers and facilitators. Multivariable regressions explored predictors of EPT support.

**Results:** Forty-eight of 70 (69%) medical directors responded, representing EDs with a median volume of 67,840 patients/year. Awareness of EPT was high (73%), but fewer knew how to prescribe it (38%), and only 19% of EDs had implemented EPT. Most (79%) supported EPT and were more likely to if they were aware of EPT (89% vs. 54%) p=0.01. Of non-implementers, 41% thought EPT was feasible, and 56% thought departmental support would be likely. Of potential barriers, ED directors were most concerned about legal liability (25% moderately to extremely). Benefits of EPT were rated with similar importance, with preventing sequelae of untreated STIs most frequently rated as “extremely important” (44%). Linear regression showed increased years in practice, and ED’s proportion of Medicaid patients was significantly positively associated with support for EPT.

**Conclusion:** ED medical directors expressed strong support for EPT and reasonable levels of feasibility for implementation but low utilization. Our findings highlight the need to identify mechanisms for EPT implementation and develop ED-tailored implementation tools to bolster this practice.

## INTRODUCTION

Rates of sexually transmitted infections (STIs) in the United States have increased considerably in the past decade.^1^ Although cases of chlamydia infection dropped slightly during the first year of the COVID-19 pandemic, cases of gonorrhea and syphilis continue to rise, contributing to a total of 2.4 million cases of STIs in 2020.^2^ As funding for sexual health clinics has decreased,^3^ emergency departments (EDs) have increasingly become a site of STI care, outpacing general ED visits.^4^ Patients at highest risk for STIs are more likely to rely on the ED for care,^5–8^ and are disproportionately from historically marginalized groups – those who are non-White, have Medicaid, or are uninsured.^9–12^

For the most common STIs, chlamydia, and most cases of gonorrhea infections, patients usually have no public health or health system assistance in ensuring their partner receives treatment.^13, 14^ However, under this current approach of patient referral – relying on the patients alone to notify their sex partner– partners are often not notified or treated,^14, 15^ exposing the patient to reinfection or recurring infection.

A strategy to prevent a patient’s reinfection from STIs is expedited partner therapy (EPT). EPT is the practice of treating the sex partners of patients with STIs – chlamydia, gonorrhea, and trichomoniasis–without an exam of the partner.^16^ It is recommended by the Centers for Disease Control and Prevention (CDC) when the partner is unlikely to receive timely care as a harm-reduction approach.^16^ Meta-analysis and systematic review of randomized control trials have shown EPT to be effective in reducing chlamydia reinfection and increasing partner treatment compared to unassisted patient referrals.^17, 18^ Despite the potential for EPT to aid in the STI epidemic via the ED – a care setting of especially high patient need– little is known about current EPT use in adult EDs at the national level. Thus, we conduct a national survey of academic ED medical directors evaluating the knowledge, attitudes, and practices regarding EPT use, as well as an examination of barriers and facilitators to its practice. Our primary outcomes were medical directors’ awareness and support of EPT and whether EPT is used in their department. We further examine how ED medical director support for EPT varies by their level of EPT awareness, hypothesizing that increasing awareness is linked with increased support. We also investigate how ED medical director EPT awareness varies by state and ED characteristics, hypothesizing that factors suggestive of an increased patient need for EPT–higher state chlamydia rates, earlier year of the state adopting EPT laws, higher ED Medicaid payer population, and patient volume– would be associated with increased EPT awareness. As an exploratory aim, we assessed whether ED medical director demographics or ED characteristics were associated with support for EPT.

## METHODS

This cross-sectional online survey was emailed to ED medical directors or emergency physicians in similar operational leadership positions using the Academy of Academic Administrators of Emergency Medicine (AAEM)/ Association of Academic Chairs of Emergency Medicine (AACEM) distribution list. Where multiple sites were listed per academic department, we used the site, the primary teaching ED for the emergency medicine (EM) residency. The AAEM/ AACEM group maintains a Benchmarking survey collecting departmental characteristics such as ED volume and proportion of patients using Medicaid insurance. We linked these benchmarking survey departmental variables to the individual responses by the ED site reported.

### SURVEY DISTRIBUTION

The survey invitation was emailed three times between July 17, 2020, to September 27, 2020. Respondents were randomized to one of two incentive levels on the first invitation round: one Amazon gift card worth $20 or a raffle to win one Amazon gift card worth $100. Randomization was conducted to assess a response rate difference between groups, for which the later wave invitations would be invited using the higher response rate incentive. Since there was no considerable difference between response rates after the first wave of distribution, the incentive was changed to a $100 gift card raffle for the remaining two email invitations. Email subject lines were descriptive as follows: “[Time – Sensitive] the <<blinded for review>> EPT Research Study” and “<<blinded for review>> Research Study: Interview for Expedited Partner Therapy.” Non-responders were followed up by emailing and calling department administrators and research faculty at the institution using publicly available email addresses, requesting that the administrator or faculty aid in requesting a medical director’s response. Survey responses were anonymous. The <<blinded for review>> institutional review board reviewed and approved the study. This study is reported following the Strengthening of the Reporting of Observational Studies in Epidemiology (STROBE) statement^19^ and previously established guidelines for EM survey research.^20^

### SURVEY DEVELOPMENT

We developed a 21-item survey instrument (**Appendix X**) including the following sections: demographics, knowledge, attitudes, and behaviors, followed by barriers and facilitators of EPT implementation. The survey instrument was created following a published framework for developing questionnaires ^21^ and began with discussions with EPT content experts alongside a literature review of published manuscripts on EPT implementation in other practice settings,^22–25^ after which new questions were developed as necessary. We refined the design and content in an iterative process—editing survey versions as needed after each step —through the following steps: (1) a discussion with a survey methodologist expert; (2) a cognitive interview with one ED medical director assessing for clarity and comprehension using the ‘think-aloud’^21^ approach, in which the interviewee verbalized his interpretations of the questions; and (3) pilot testing with one other ED medical director who provided written feedback to correct any technical issues and ensure questions were appropriate, and question options were complete. The survey was designed to take under 5 minutes.

### SURVEY CONTENT

The primary outcomes were EPT awareness, support, and practices. Awareness was assessed by knowledge of EPT’s definition, departmental to state-level guidelines, and STI-specific indications. Medical directors’ support, as well as their perception of support from other ED stakeholders—other clinician prescribers, nurses, and the ED as a department– were rated by the medical director on a 5-point Likert scale (1= “Strongly Oppose”; 5= “Strongly Support”; “Unsure”). Perception of departmental support was asked on a scale assessing likelihood (1= “Extremely unlikely”; 5= “Extremely likely.”) Perceived feasibility for instituting EPT was assessed on a 4-point Likert scale (1=“Somewhat Feasible,” 4= “Very Feasible”; “Unsure”). For past practices, respondents were asked if they had personally prescribed EPT and if it was implemented at their ED.

We further analyzed whether EPT awareness varied by pre-determined state-level variables (chlamydia rates, year of adopting EPT laws) and departmental-level variables (proportion of Medicaid-insured patients, ED annual volume). For state variables, based on prior literature,^25^ we examined early versus late adopters of EPT permissible laws as well as state-level chlamydia rates. EPT law adoption status was determined based on a combination of state data pre-classified from previous research^25^ and updated according to the CDC.^26^ Early adopters were classified as states with EPT laws from 2001-2014, and late adopters were states with EPT laws from 2015-2019. Chlamydia rates were based on data from the CDC^27^ and states were categorized based on if their rate was above or below the 2015 median case rate of 447 cases per 100,000 people. For departmental variables, we examined data from the AAEM/ AACEM Benchmarking survey of ED volume and the proportion of Medicaid insurance patients. Survey invitation links were associated with an institution-specific code to allow linkage of known state and departmental variables.

To assess barriers and facilitators of EPT, respondents first rated their level of concern for potential barriers on a 5-point Likert scale (1= “Not at all concerned”; 2= “Slightly”; 3= “Concerned”; 4=” Moderately concerned”; 5= “Extremely concerned”; “Unsure”) for the following topics: adverse reaction, missed diagnoses, concern for intimate partner violence, legal liabilities, affordability of EPT medications, and the ability for the pharmacy to fill prescriptions. Respondents then rated the importance of potential benefits on a 5-point Likert similar to above, instead using the keyword “important” in place of “concerned” for the following topics: preventing STI reinfection, increasing access to treatment among vulnerable populations, addressing untreated STIs, and preventing sequela of STIs.

### STATISTICAL ANALYSIS

Descriptive statistics included proportions with 95% confidence intervals to report demographic characteristics and main outcome variables. We conducted a bivariate analysis using the Fisher exact test to assess whether awareness of EPT (“Yes” vs. “No”) was associated with EPT attitudes and practice or departmental and state characteristics. The top-box approach was used for the analysis of barriers and facilitators. The “top-box” score indicates the proportion of respondents who selected the highest response category and is commonly used by hospital patient experience surveys.^28, 29^

As an exploratory analysis, we used multivariable linear regression tests to evaluate predictors of EPT support. We used a theory-driven approach to select covariates: state-level^25^ (chlamydia prevalence and onset of EPT laws), ED-level (ED volume and proportion of patients using Medicaid insurance), and respondent-level (gender and years in practice). For the outcome of the medical director’s perception of departmental support, we added the following covariates: perceived support from prescribers and RN, feasibility, and awareness of hospital policies.

The response rate was calculated via American Association for Public Opinion Research (AAPOR) response rate 1 definition, which considers surveys at least 80% complete divided by the total contacted.^30^ As a non-response bias analysis,^31^ respondents were compared to nonrespondents by available ED sites and respondent characteristics. The survey was administered via Qualtrics (Provo, Utah) and data was analyzed via R statistical computing (Vienna, Austria) for analysis. A p-value ≤0.05 was considered statistically significant.

## RESULTS

Of 70 potential respondents, 48 submitted complete surveys (AAPOR response rate of 69%, 100% completion rate, no incomplete or break-offs); demographics, ED, and state characteristics are described in **Table 1**. Respondents were 31% female, and more than half (52%) had over five years in their leadership roles. Surveyed EDs were geographically distributed across the country, with most from the Northeast (33%). Most (79%) EDs were in states that were early adopters of EPT laws (between 2001-2014). Under half (42%) were in states with high chlamydial incidence. Comparing non-responders to responders, we observed no significant differences across key characteristics for which data were available: medical director gender, ED region, volume, percentage Medicaid, EPT adoption year, and chlamydia incidence (**eTable 1).**

**Table 1.** Demographic and ED Characteristics of Respondents

|  | Total No | Response (%) |
| --- | --- | --- |
| Med Director Characteristic | (n=48) |  |
| Female | 15 | 31 |
| Year in practice |  |  |
| 0-5 years | 1 | 2 |
| 6-10 years | 12 | 25 |
| 11-20 years | 24 | 50 |
| 21+ years | 11 | 23 |
| Year in role |  |  |
| 0-5 years | 23 | 48 |
| 6-10 years | 14 | 29 |
| 11-15 years | 7 | 15 |
| 16+ year | 4 | 8 |
| **ED characteristic\*** |  |  |
| Region |  |  |
| Midwest | 8 | 17 |
| Northeast | 16 | 33 |
| South | 14 | 29 |
| West | 10 | 21 |
| Medicaid |  |  |
| Mean |  | 28 |
| Median |  | 26 |
| IQR |  | 20 |
| ED Volume |  |  |
| Mean |  | 74,699 |
| Median |  | 67,840 |
| IQR |  | 25,183 |
| **State characteristic\*\*** |  |  |
| Timing of EPT laws |  |  |
| Early adopter | 38 | 79 |
| Later adopter | 10 | 21 |
| Chlamydia incidence |  |  |
| Low incidence | 28 | 58 |
| High incidence | 20 | 42 |

Note: States law timing is classified as follows: earlier adopters (2001-2014) or late (2015-2019). Chlamydia incidence is classified as follows: low (233-445/100K population) or high(455-768/100K population)

### EPT AWARENESS

Most medical directors (73%) knew of EPT (**Table 2**). Of those reportedly aware of EPT, most (66%) could correctly identify that EPT was used for chlamydia in their state. Less than half of medical directors were aware of the existence of guidelines from the CDC (48%), and fewer were aware of more local-level guidance: State law (31%), local health department (21%), and local hospital (15%). A little over a third of medical directors reported that they knew how to write a prescription (38%), and fewer had previously prescribed (31%). Only 19% reported that their ED had implemented EPT at the departmental level.

**Table 2.**
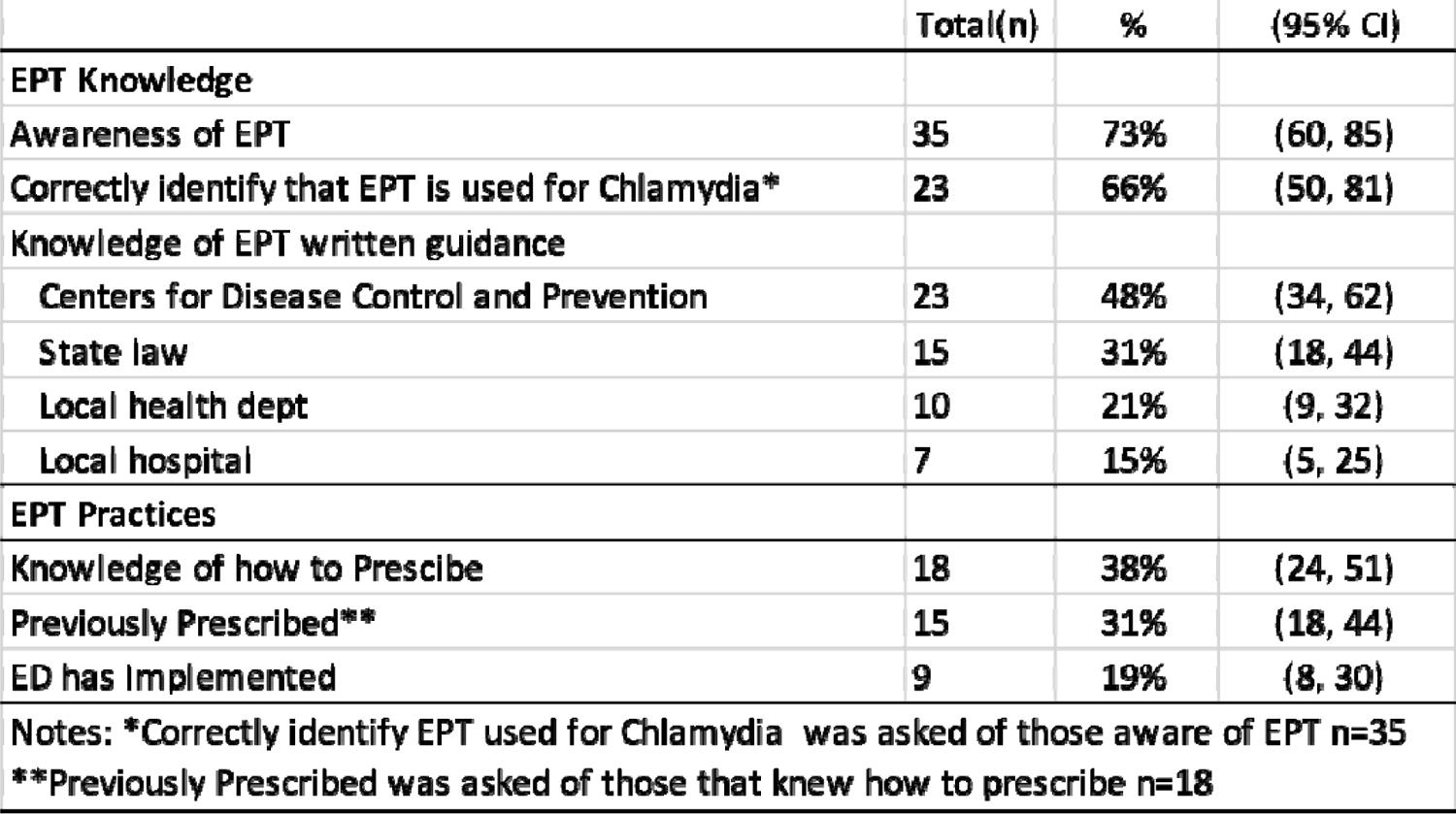
Emergency Department Medical Director Knowledge and implementation of Expedited Partner Therapy

|  |  |  |  |
| --- | --- | --- | --- |
|  | **Total(n)** | **%** | **(95% CI)** |
| **EPT Knowledge** |  |  |  |
| Awareness of EPT | 35 | 73% | (60, 85) |
| Correctly Identify that EPT is used for Chlamydia* | 23 | 66% | (50, 81) |
| **Knowledge of EPT written guidance** |  |  |  |
| Centers for Disease Control and Prevention | 23 | 48% | (34, 62) |
| State law | 15 | 31% | (18, 44) |
| Local health dept | 10 | 21% | (9, 32) |
| Local hospital | 7 | 15% | (5, 25) |
| **EPT Practices** |  |  |  |
| Knowledge of how to Prescribe | 18 | 38% | (24, 51) |
| Previously Prescribed** | 15 | 31% | (18, 44) |
| ED has Implemented | 9 | 19% | (8, 30) |
| Notes: *Correctly identify EPT used for Chlamydia was asked of those aware of EPT n=35 |  |  |  |
| **Previously Prescribed was asked of those that knew how to prescribe n=18 |  |  |  |

### EPT SUPPORT

**Table 3** shows medical director support and perceived institutional support of EPT stratified by awareness level of EPT. There were high support from medical directors towards EPT (79%) and a perception of similarly high support from other prescribers (71%). Conversely, respondents felt nurses would have lower support for EPT (50%). Support was significantly higher when respondents were aware of EPT both for medical director self-reported support (P<0.001) and their perception of nurse support (P<0.04). Regarding departmental factors, 41% of participants expected the implementation of EPT at their site to be “very feasible”(18%) or “feasible”(23%). Only 5% reported it was “not at all feasible.” Over half (56%) of medical directors anticipated that their ED would be supportive of EPT at either “Somewhat likely” (49%) or “Extremely likely” (8%) levels. Approximately two-thirds (67%) of medical directors felt they would be impacted in their perception of EPT if peer institutions were already implementing it.

**Table 3.**
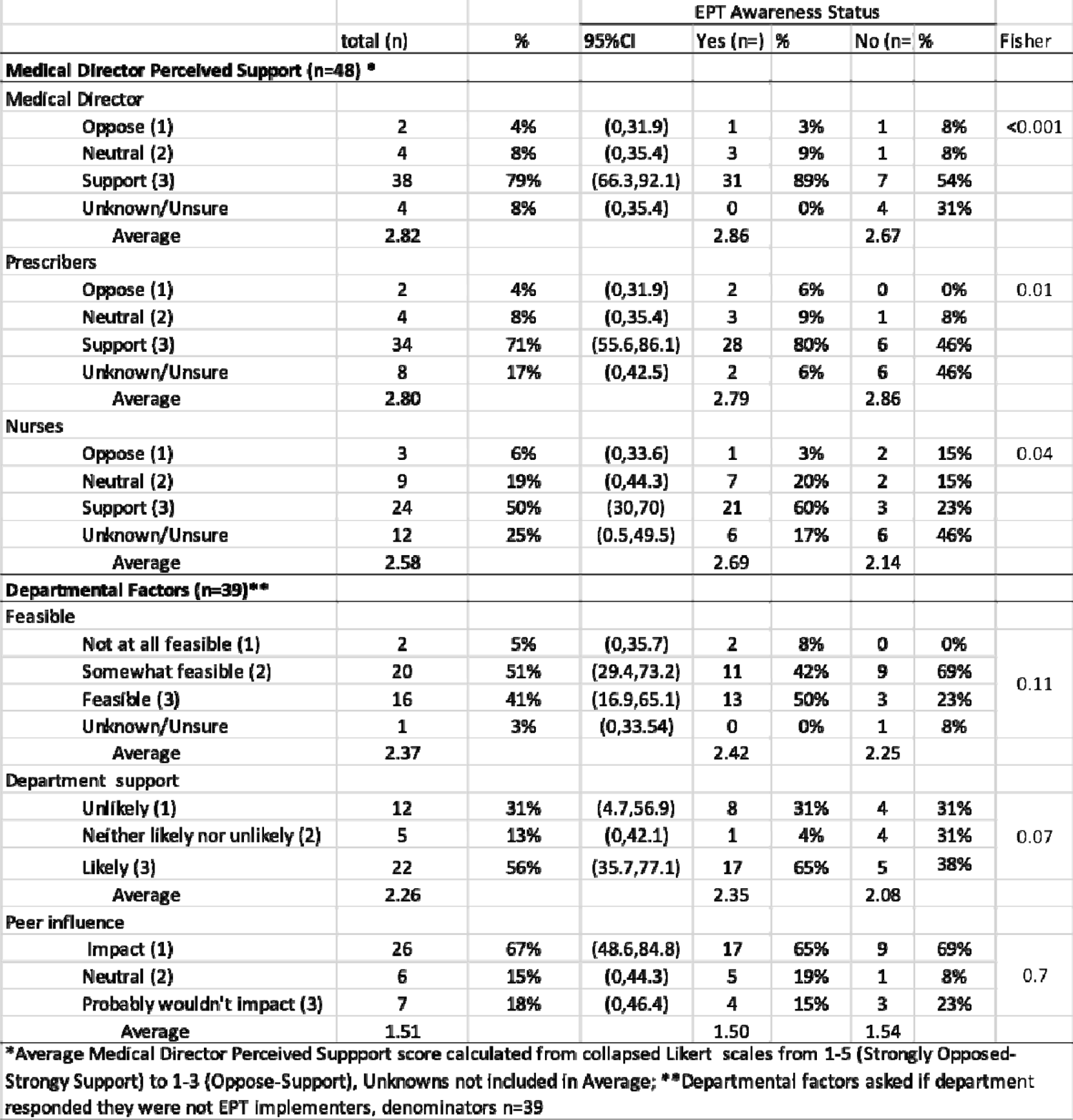
Medical Director Attitudes and Perceptions of Expedited Partner Therapy

### VARIATION WITH STATE AND ED CHARACTERISTICS

We examined whether medical director awareness of EPT varied by state or departmental characteristics (**eTable 2).** Respondent awareness of EPT varied significantly by state chlamydia incidence level (Fisher exact test p< 0.05), such that in states with higher chlamydia incidence, most respondents (90%, 18/20) were aware of EPT and most respondents (85%, 11/13) unaware of EPT were from lower chlamydia incidence states. Other potential characteristics had no associated difference in EPT awareness status: ED volume, the proportion of Medicaid patients, and the year when state adopted laws supporting EPT.

### BARRIERS AND FACILITATORS TO EPT

The EPT barriers and facilitators showed only slight variation in their ranking. The perceived barriers to EPT are presented ranked in order of highest percentage of most concerned **(eFigure 1, tabular form in eTable 3)**: (1) legal liability (25%), (2) intimate partner violence (21%), (2) affordability of EPT medication (21%), (3) potential missed medical diagnoses (19%), (4) ability for the pharmacy to fill prescriptions (17%), (5) adverse reactions to the antibiotics (13%). The highest ranked benefits to patients were: (1) increasing access to treatment among vulnerable populations (81%), (1) addressing untreated STIs (81%), (1) preventing sequela of STIs (81%), and (2) preventing STI reinfection (79%). There were minimal differences in the mean scores between barriers and facilitators. Overall, many more respondents rated facilitators at the highest level of importance compared to the proportion that rated barriers with the highest level of concern: 81% rated the top three facilitators as the highest importance compared to 25% for the top barrier.

### PREDICTORS OF EPT SUPPORT

Exploratory regression analysis in variables associated with increasing support for EPT found the following were significantly related: years in practice, the proportion of patients using Medicaid, and the feasibility level of EPT. For the outcome of medical director support (Strongly Opposed (1)- Strongly Support (5)), every unit increase in years practicing emergency medicine (0-5 years; 6-10 years; 11-20 years; over 21 years), corresponded to a 1.02 point (95%CI 0.6, 1.44) p<0.0001 increase in the level of EPT support. Conversely, every unit increase in years in the medical director role (0-5 years; 6-10 years; 11-15 years; over 16 years) corresponded to a −0.7 point (95%CI −1.06, −0.34) P<0.001 decrease in support. For every 10% increase in the proportion of Medicaid patients, there was a 0.24 point (95%CI 0.04,2.37) p=0.04 increase in support. For the outcome of departmental EPT support, for every increase in perceived feasibility of EPT (Not at all feasible (1)-Very feasible (4)) there was a 0.7 (95%CI 0.24-1.13) p=0.01 increase in support.

## DISCUSSION

This study represents the first national evaluation of knowledge and support for EPT among ED medical directors. While most medical directors were aware of EPT, only some knew how to prescribe it, and even fewer had written prescriptions. Together with their high ranking of the benefits of EPT, this gap between interest and practice highlights the role that more macro-level factors, such as institutional or cultural, may play in the low use of EPT. Moreover, compared to their stated high support levels, medical directors’ perception of comparatively lower support from their ED, other healthcare providers, and nurses suggests a tension between personal perspectives on patient care and organizational realities.

Potential explanations for this implementation gap may be due to medical directors’ perceptions of barriers regarding legal liability, the potential for misdiagnosis, affordability of medication, and intimate partner violence. Despite concern for these barriers, medical directors still espoused the importance of EPT benefits, including increasing access to treatment among vulnerable populations, preventing STI reinfection, preventing sequela of STIs, and addressing untreated STIs.

Legal concerns remain of high importance to medical directors. Despite EPT ^26^ being permissible or potentially allowable in all U.S. states, 25% of ED directors stated that legal concerns were an “extreme concern.” This concern may be partly due to the unfamiliarity of EPTs’ prescribing practice to an unseen individual and lack of familiarity with institutional policies to support ETP’s use-only 15% knew of hospital-level policies. There has been little to no national research on this topic in adult EDs. This unfamiliarity with EPT in practice and policy likely contributes to decreased comfort with EPT as a practice option. This may change in coming years as ED organizations have adopted policies supporting the use of EPT, such as the American College of Emergency Physicians’ policy supporting the development of EPT protocols and model state legislation to remove legal obstacles to EPT.^32^ Importantly, in many states, there are already explicit liability protections for the provider to use EPT, and there have been no reported medical malpractice cases involving EPT.^33^ Additionally, the safety of EPT has been well established: No adverse reactions were recorded in patients included in randomized control trials,^17^ nor the fifteen years during which a hotline to report adverse events was open in California.^34^ Increased awareness of supportive policies, legal protections, and patient preferences may help alleviate some of the medical director’s concerns.

Past work has similarly identified low levels of EPT use among healthcare providers, particularly in EM. A survey of physician members of the American Academy of Pediatrics Section of Emergency Medicine found that only 30% of physicians were aware of state EPT laws,^35^ similar to our finding that 31% stated awareness of their state laws. A study that analyzed data on gonorrhea cases from 12 sites in the CDC’s STD Surveillance Network found that only 10% of patients with gonorrhea reported receiving EPT for their partners.^36^ Previous surveys comparing EPT across specialties found that EPT uptake in the ED was lower than in other specialties-most (56%) of ED physicians had never used partner therapy, and only 13% used partner therapy “half or more” of the time.^37^ One study of 492 patients within the Indian Health Service found lower rates of EPT use in urgent care or EDs compared to outpatient settings.^38^ In a survey of a convenience sample of healthcare providers in Pittsburg, PA, from diverse disciplines who treat young women at risk for chlamydia, only 11% of providers were using EPT consistently.^22^ In contrast to our finding of 19% of EDs reporting EPT use, a study of an urban, multicenter safety net institution in Georgia found that while ED providers were willing to consider EPT, none were actually practicing it. This study also identified unique barriers to antibiotic overuse in presumptive or symptomatic treatment without laboratory confirmation, as well as concern for increasing traffic to the ED for STI patients.^23, 24^

### LIMITATIONS

Our study has limitations. This study had a relatively small sample size, and we only surveyed medical directors in academic EDs. These results may not be generalizable to a broader spectrum of emergency departments, especially those in a community setting. While we use state-level chlamydia infection rates as a potential predictor of EPT support and awareness, state-level rates of STI infections may not reflect local prevalence or community resources, which are more likely to influence ED practices. Moreover, we only included medical directors due to the administrative and operations focus of instituting new departmental workflows and policies. But medical directors may have incomplete knowledge of EPT feasibility due to unfamiliarity with specific legal liability protections. Their support for EPT may be less valuable for the implementation of the process than a departmental physician champion who could take ownership of the process. Also, the director’s perceptions of the support of other providers, such as nurses, is a perception and not the actual perspective of other providers. Like survey work generally, the group of directors responding to the survey invitation is susceptible to response bias, where ED directors more interested in reproductive health may be more likely to respond, resulting in an overrepresentation of EPT awareness, practice, and support.

Additionally, survey collection occurred during the COVID-19 pandemic, which caused significant administrative stressors on EDs, especially medical directors. This would have limited their ability to respond to a voluntary survey, further impacting the selection of respondents. Survey questions may have been susceptible to social desirability bias since the target EPT population has lower healthcare access. Thus, supporting practices to assist this population may be perceived as more socially acceptable.

Strengths of the study included a high overall response rate (69%) and the inclusion of EDs from a range of US geographical areas and areas with varying STI prevalence. We analyzed EPT across important metrics such as the proportion of Medicaid patients and ED volume using SAEM Benchmarking survey group data. To investigate response bias, we compared these ED metrics and geographical variables of STI prevalence and recency of EPT state law adoption. We observed no statistically significant differences between respondents to non-respondents.

The use of EPT in the high-need setting of the ED is increasingly crucial as STI cases increase and health disparities in STIs remain a problem of health equity. Social determinants of health impact STI epidemiology, and there is an extensive history of STI disparities associated with gender, sexual orientation, age, income, and race/ ethnicity. ^6, 9, 12, 39–43^ According to the CDC, in 2019, gonorrhea rates were 42 times higher among men who have sex with men and bisexual men compared to heterosexual men.^44^ Young women account for 43% of reported cases of chlamydia and risk severe consequences such as pelvic inflammatory disease and infertility.^45^ Moreover, undiagnosed STIs may contribute to infertility in over 20,000 women annually.^44^ The ED’s awareness of these disparities and increased use of strategies such as EPT could help combat worse sexual and reproductive health outcomes among historically socially disadvantaged populations. Future research should further characterize barriers and facilitators to ED-EPT implementation in the ED in greater detail. A qualitative analysis of best practices where EPT is conducted will lead to a better understanding of how interested EDs can implement EPT.

## CONCLUSION

A national study of academic EDs found that 79% of medical directors supported EPT; however, only 19% reported that their department had implemented EPT, indicating a significant opportunity to increase the adoption of this evidence-based practice. The ED can play a critical public health role in stemming the spread of curable STIs, disproportionately affecting historically marginalized populations.

## Data Availability

All data produced in the present study are available upon reasonable request to the authors

## ACKNOWLEDGMENTS

The study team thanks <<<Blinded>> survey expert who helped review the survey instrument and provided valuable feedback.

## SUPPLEMENTAL MATERIAL

**eTable 1.**
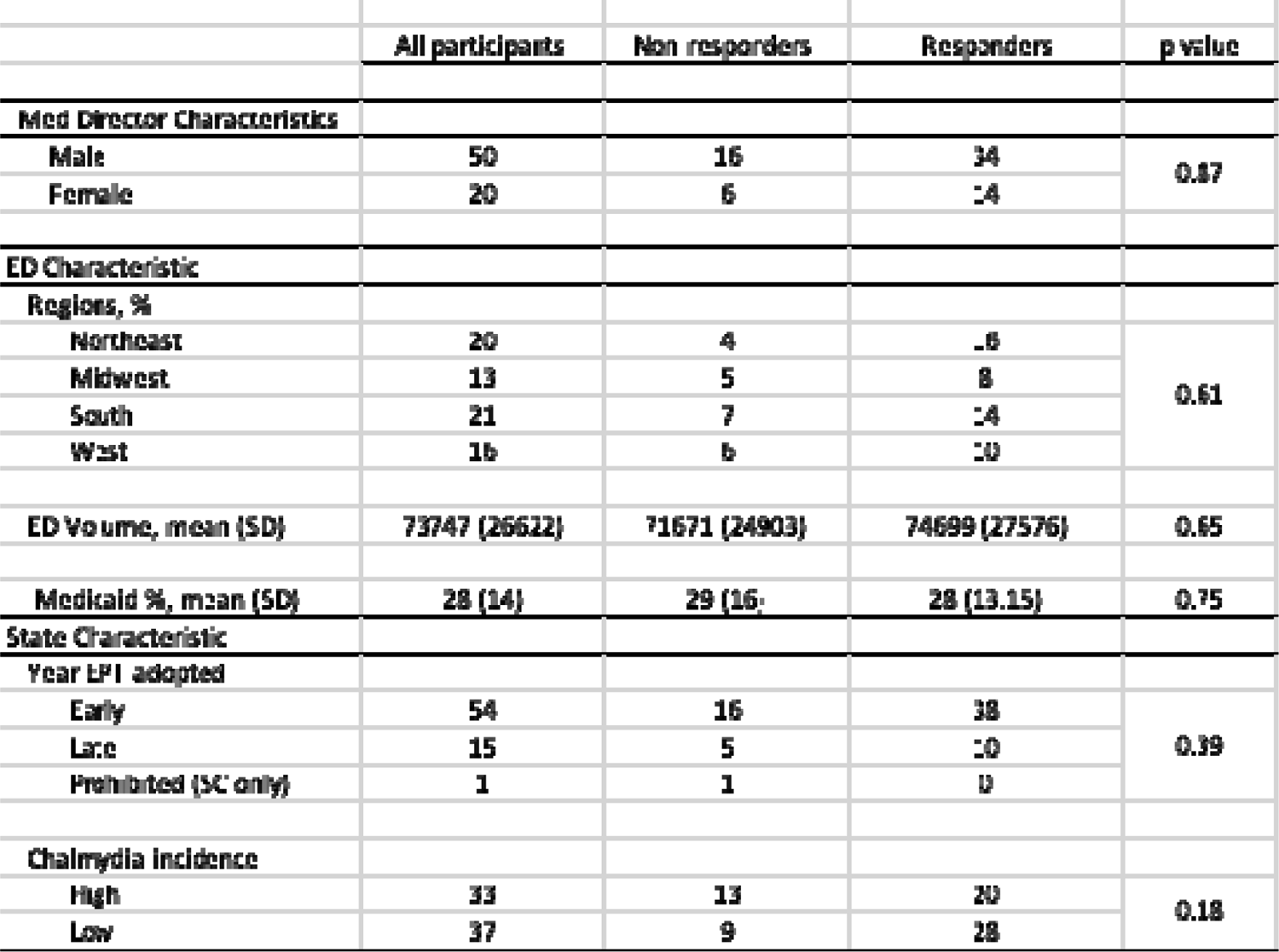
Characteristics of non-responders versus responders

**eTable 2.**
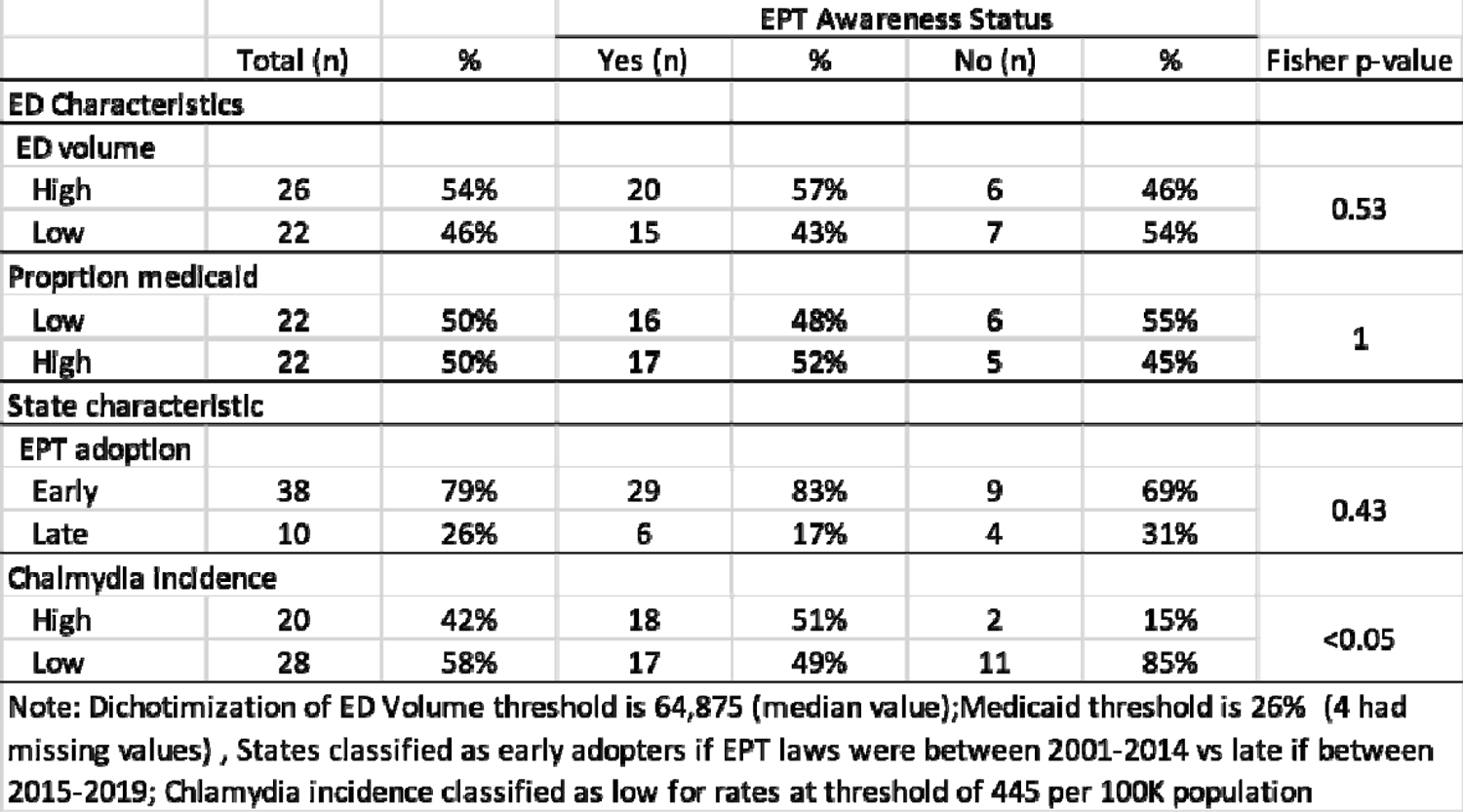
Expedited Partner Therapy Awareness by Emergency Department and State Characteristics

**eFigure 1.**
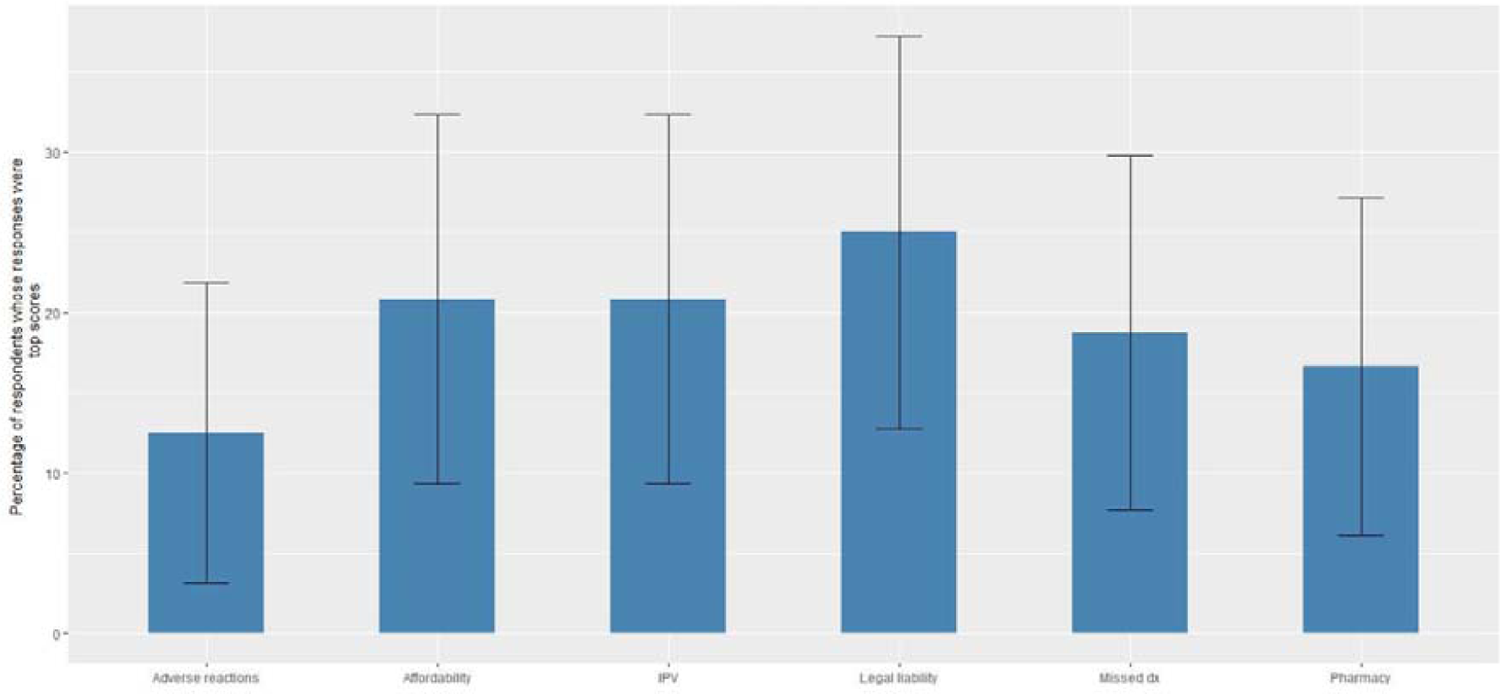
Medical Director Perceived Barriers to Expedited Partner Therapy

**eTable 3.**
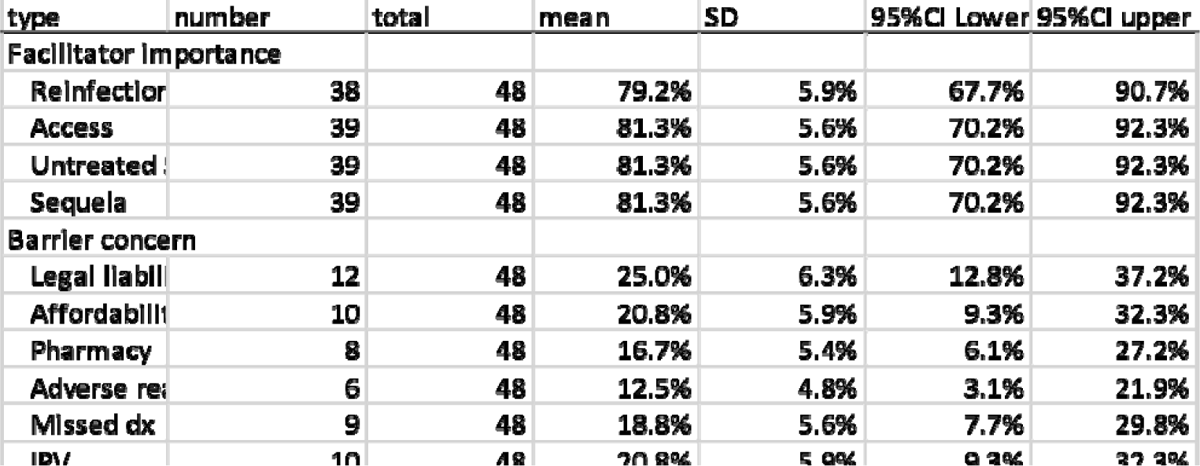
Percentage of medical directors reporting highest rating of concern or Importance for barriers and facillitators of expedited partner therapy

## SURVEY INSTRUMENT

consent Expedited Partner Therapy Survey This survey is intended for the Medical Director of Emergency Departments (ED) or physicians in similar operational leadership roles. We are a team of emergency physicians and researchers from <<blinded for review>> studying how EDs treat patients and their patients’ sexual partners for STIs. We also are studying Medical Directors’ knowledge and perspectives on treating patients’ sexual partners and the clinical practice of Expedited Partner Therapy (EPT). If you are unfamiliar with EPT we will explain it later in the survey. The survey should take 5 minutes. Answering this survey is voluntary. You can skip any questions that you don’t want to answer. Answering our survey won’t benefit you directly. What we learn will help us in guiding the implementation of policies that increase access to STI care. To keep your information confidential, your survey responses will be anonymous. For questions, please contact <<blinded for review>> MD and the research team at <<blinded for review>>. As a thank you for participation, you will have the option to receive a $20 Amazon gift card or to contribute the money back to EPT research at the survey’s completion. If you would like to enter the drawing, you can anonymously enter your email after the survey is completed. Your email will not be stored with your survey responses. To participate, check the “I agree to participate” box below, then click the blue button to begin.

- Yes, I agree to participate (1)
- No, I do not agree to participate (0)

End of Block: Consent

Start of Block: Demographics / Who are you

role Which primary operational leadership role best describes your position in the emergency department?

- Medical Director of the Emergency Department (1)
- Division Chief of Emergency Medicine (2)
- Service Chief of Emergency Medicine (3)
- Chief of Staff (4)
- (Vice) Chair/ Director of (Clinical) Operations (5)
- (Vice) Chair/ Director of Quality and Safety (6)
- (Vice) Chair/ Director of Clinical and Faculty Affairs (7)
- Clinical Director (8)
- Other (9)

years_EM How many years have you been practicing emergency medicine?

- 0-5 years (1)
- 6-10 years (2)
- 11-20 years (3)
- 21+ years (4)

years_role How many years have you been in an operational leadership role in the ED?

- 0-5 years (1)
- 6-10 years (2)
- 11-15 years (3)
- 16+ years (4)

End of Block: Demographics / Who are you

Start of Block: Knowledge

EPTaware Expedited Partner Therapy (EPT) is the clinical practice of treating the sexual partners of patients diagnosed with chlamydia or gonorrhea (and/ or trichomonas) by providing prescriptions or medications to the patient to take to his/ her partner(s) *without the health care provider first examining the partner.* This practice is also sometimes alternatively called Patient-Delivered (Partner) Therapy. Have you ever heard about Expedited Partner Therapy (EPT) before?

- Yes (1)
- No (0)

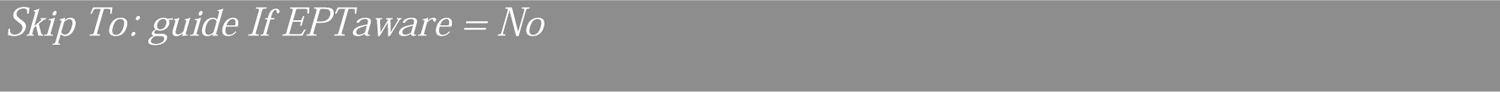

STI To the best of your knowledge, which STIs do you think can be treated with EPT in your state?

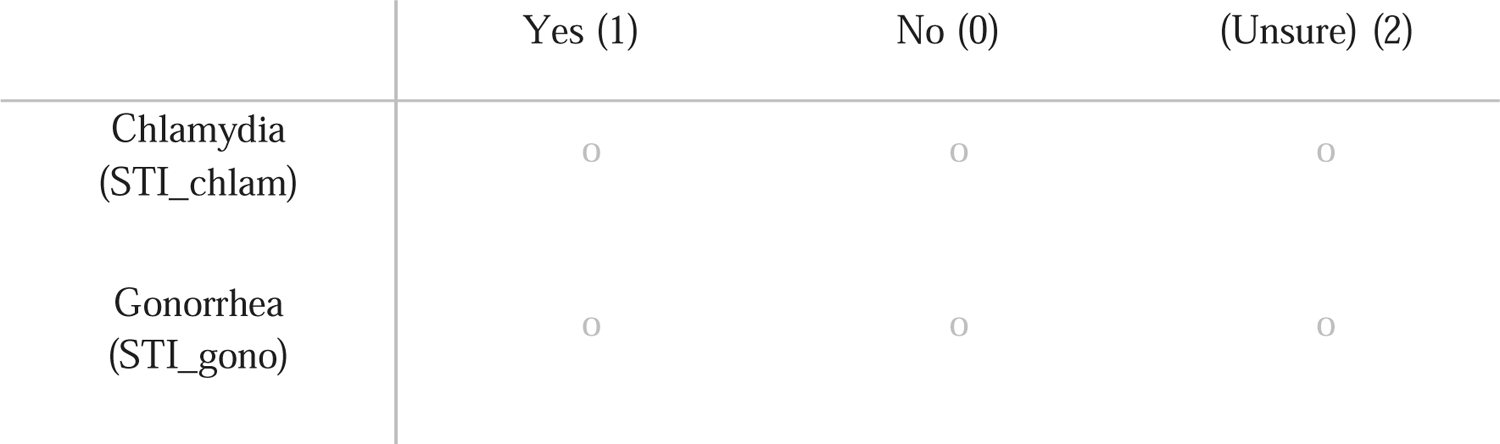
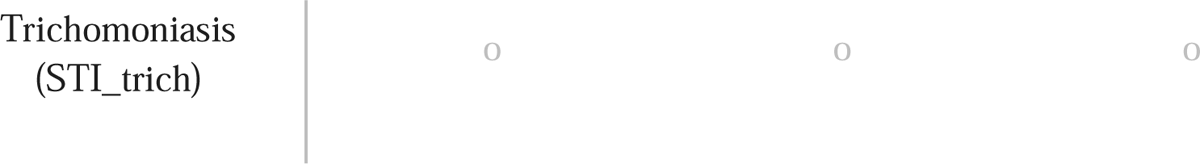

guide To the best of your knowledge, which of the following groups have written guidance regarding EPT?

|  | Yes (1) | No (0) | (Unsure/ Don't know) (2) |
| --- | --- | --- | --- |
| CDC guidelines<br>(guide_CDC) | ☐ | ☐ | ☐ |
| State laws<br>(guide_law) | ☐ | ☐ | ☐ |
| Local health<br>department<br>(guide_local) | ☐ | ☐ | ☐ |
| Local hospital<br>guidelines<br>(guide_hospital) | ☐ | ☐ | ☐ |

End of Block: Knowledge

Start of Block: Interest and Feasibility

support In your opinion, rate the level of support or expected level of support of the following ED clinicians for EPT being offered at your facility?

|  | Strongly<br>Oppose<br>(1) | Somewhat<br>Oppose<br>(2) | Neutral<br>(3) | Somewhat<br>Support<br>(4) | Strongly<br>Support<br>(5) | (Unsure<br>) (6) |
| --- | --- | --- | --- | --- | --- | --- |
| Your personal<br>interest<br>(support_meddir) | ☐ | ☐ | ☐ | ☐ | ☐ | ☐ |
| Physicians/<br>Physician<br>Assistants (PA) /<br>Nurse Practitioners<br>(NP)<br>(support_prescriber<br>) | ☐ | ☐ | ☐ | ☐ | ☐ | ☐ |
| Nurses (RN)<br>(support_RN) | ☐ | ☐ | ☐ | ☐ | ☐ | ☐ |

implementer Has your emergency department already implemented EPT? (i.e. Are physicians/ PAs/ NPs already writing prescriptions for the sexual partners of patients diagnosed with STIs without an exam of the partner?)

- Yes (1)
- No (0)
- (Unsure) (2)

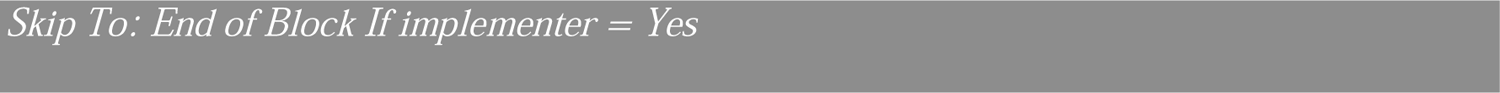

feasible How feasible do you think it would be to implement EPT at your ED?

- Not at all feasible (1)
- Somewhat feasible (2)
- Feasible (3)
- Very feasible (4)
- (Unsure) (5)

departsupport Do you think your department would support implementing EPT, including clinical staff time for patient follow-up and counseling?

- Extremely unlikely (1)
- Somewhat unlikely (2)
- Neither likely nor unlikely (3)
- Somewhat likely (4)
- Extremely likely (5)

peers Would EPT being practiced at peer institutions have an impact on your decision to implement EPT at your ED?

- Definitely would impact (1)
- Probably would impact (2)
- Neutral (3)
- Probably would not impact (4)
- Definitely would not impact (5)

End of Block: Interest and Feasibility

Start of Block: Prior Experience

writerx Do you know how to write a prescription for EPT?

- Yes (1)
- No (0)

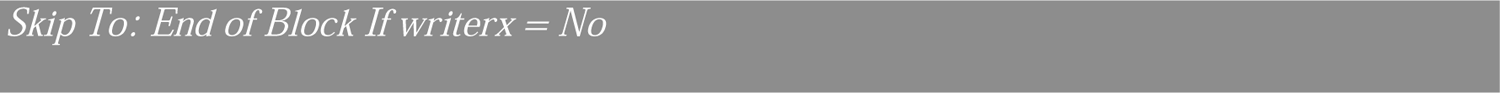

prescribed

Have you prescribed medications through EPT?

- Yes (1)
- No (0)

End of Block: Prior Experience

Start of Block: Risks and Benefits

concernLOG Rate your level of concern regarding the following potential logistical challenges with EPT.

|  | Not at all<br>concerned (1) | Slightly<br>concerned (2) | Concerned (3) | Moderately<br>concerned (4) | Extremely<br>concerned (5) | (Unsure) (6) |
| --- | --- | --- | --- | --- | --- | --- |
| Legal liability concerns<br>(concernLOG_legal) | ○ | ○ | ○ | ○ | ○ | ○ |
| Affordability of EPT medication<br>(concernLOG_afford) | ○ | ○ | ○ | ○ | ○ | ○ |
| Ability for pharmacy to fill Rx<br>(concernLOG_pharm) | ○ | ○ | ○ | ○ | ○ | ○ |

concernPT Rate your level of concern regarding the following potential patient outcome issues with EPT.

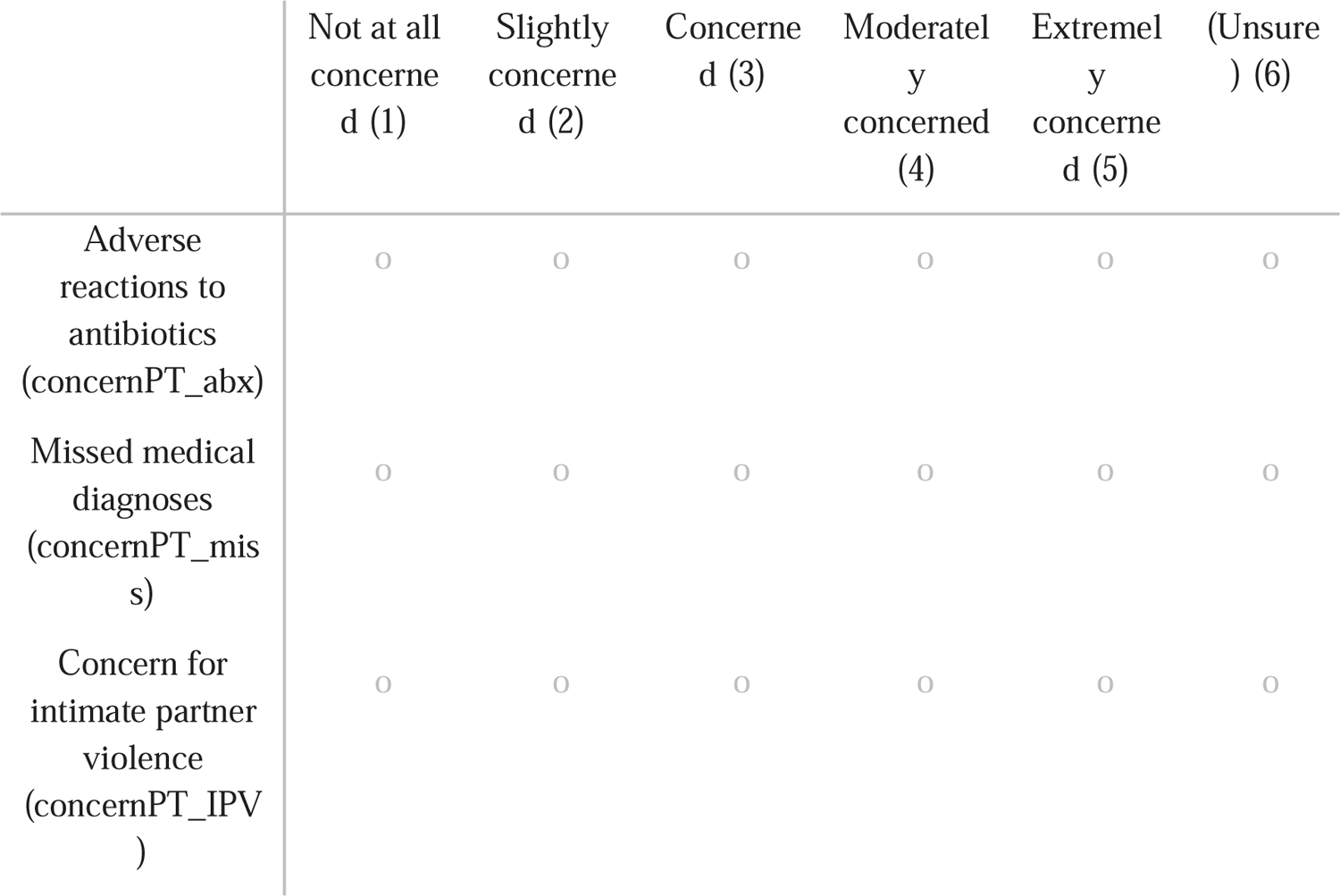

concern_freetext Do you have additional concerns about potential risks with EPT?

benefit In your opinion, rate the importance of the following potential benefits with EPT:

concern\_fretext Do you have additional concerns about potential risks with EPT? benefit In your opinion, rate the importance of the following potential benefits with EPT:
|  | Not<br>important<br>(1) | Slightly<br>important<br>(2) | Moderately<br>important<br>(3) | Very<br>important<br>(4) | Extremely<br>important<br>(5) | (Unsure)<br>(6) |
| --- | --- | --- | --- | --- | --- | --- |
| Preventing STI<br>reinfection<br>(benefit_reinfx) | 0 | 0 | 0 | 0 | 0 | 0 |
| Increasing<br>access to<br>treatment among<br>vulnerable<br>populations<br>(benefit_access) | 0 | 0 | 0 | 0 | 0 | 0 |
| Addressing<br>untreated STIs<br>(benefit_STI) | 0 | 0 | 0 | 0 | 0 | 0 |
| Preventing<br>sequela of STIs<br>(i.e pelvic<br>inflammatory<br>disease)<br>(benefit_prevent<br>) | 0 | 0 | 0 | 0 | 0 | 0 |

benefit_freetext Do you see any additional benefits with EPT?

End of Block: Risks and Benefits

## Notes

**Financial Support:** Dr. Solnick was supported by the U-M National Clinician Scholars Program at the Institute for Healthcare Policy and Innovation during this project. This work was also supported by the University of Michigan Resident and Fellow Research Development Grant award (RDG).

### Competing Interest Statement

The authors have declared no competing interest.

### Funding Statement

Dr. Solnick was supported by the U-M National Clinician Scholars Program at the Institute for Healthcare Policy and Innovation during this project. This work was also supported by the University of Michigan Resident and Fellow Research Development Grant award (RDG).

### Author Declarations

The University of Michigan institutional review board gave ethical approval for this work.

